# Trajectories of host-response biomarkers and inflammatory subphenotypes in COVID-19 patients across the spectrum of respiratory support

**DOI:** 10.1101/2022.11.28.22282858

**Authors:** Michael Lu, Callie Drohan, William Bain, Faraaz A. Shah, Matthew Bittner, John Evankovich, Niall Prendergast, Matthew Hensley, Tomeka Suber, Meghan Fitzpatrick, Raj Ramanan, Holt Murray, Caitlin Schaefer, Shulin Qin, Xiaohong Wang, Yingze Zhang, Seyed M. Nouraie, Heather Gentry, Cathy Kessinger, Asha Patel, Bernard J. Macatangay, Jana Jacobs, John Mellors, Janet S. Lee, Prabir Ray, Anuradha Ray, Barbara Methé, Alison Morris, Bryan J. McVerry, Georgios D. Kitsios

## Abstract

**Purpose:** Enhanced understanding of the dynamic changes in the dysregulated inflammatory response in COVID-19 may help improve patient selection and timing for immunomodulatory therapies.

**Methods:** We enrolled 323 COVID-19 inpatients on different levels of baseline respiratory support: i) Low Flow Oxygen (37%), ii) Non-Invasive Ventilation or High Flow Oxygen (NIV_HFO, 29%), iii) Invasive Mechanical Ventilation (IMV, 27%), and iv) Extracorporeal Membrane Oxygenation (ECMO, 7%). We collected plasma samples upon enrollment and days 5 and 10 to measure host-response biomarkers. We classified subjects into inflammatory subphenotypes using two validated predictive models. We examined clinical, biomarker and subphenotype trajectories and outcomes during hospitalization.

**Results:** IL-6, procalcitonin, and Angiopoietin-2 were persistently elevated in patients at higher levels of respiratory support, whereas sRAGE displayed the inverse pattern. Patients on NIV_HFO at baseline had the most dynamic clinical trajectory, with 26% eventually requiring intubation and exhibiting worse 60-day mortality than IMV patients at baseline (67% vs. 35%, p<0.0001). sRAGE levels predicted NIV failure and worse 60-day mortality for NIV_HFO patients, whereas IL-6 levels were predictive in IMV or ECMO patients. Hyper-inflammatory subjects at baseline (<10% by both models) had worse 60-day survival (p<0.0001) and 50% of them remained classified as hyper-inflammatory on follow-up sampling at 5 days post-enrollment. Receipt of combined immunomodulatory therapies (steroids and anti-IL6 agents) was associated with markedly increased IL-6 and lower Angiopoietin-2 levels (p<0.05).

**Conclusions:** Longitudinal study of systemic host responses in COVID-19 revealed substantial and predictive inter-individual variability, influenced by baseline levels of respiratory support and concurrent immunomodulatory therapies.

## Introduction

SARS-CoV-2 has infected more than 615 million individuals and led to more than 6.5 million deaths worldwide[1], with more than 1 million deaths in the USA[2] as of October 2022. Extensive research has shown that COVID-19 patients with severe illness requiring hospitalization develop a dysregulated inflammatory response against the virus, often leading to acute respiratory failure with parenchymal lung damage and impaired gas exchange[3]. Current care consists of two main elements: i) provision of appropriate respiratory support (invasive or non-invasive options) to improve gas exchange and work of breathing, and ii) delivery of timely and effective antiviral and immunomodulatory therapies[4, 5] to curtail the aberrant inflammatory response.

The provision of the first main element of care, providing appropriate respiratory support, is dynamic and responsive to clinical changes at the bedside. Provision of the second main element of care, antiviral and immunomodulatory agents, is based largely on cross-sectional assessments of respiratory failure severity and crude biomarkers that are available clinically (e.g. C-reactive protein levels for anti-IL-6 treatment initiation). However, the systemic inflammatory response in severe COVID-19 is complex, with multiple pathways involved and differences compared to non-COVID acute respiratory distress syndrome (ARDS)[6]. Extensive research in non-COVID ARDS has shown replication validity of distinct host-response subphenotypes (e.g. hyper- and hypoinflammatory), potentially offering new opportunities for targeted therapeutics[7-10]. Such biomarker-based subphenotypes have also been described in COVID-19 ARDS and may allow better targeting of immunomodulatory interventions. Enhanced understanding of the dynamic variability of the longitudinal systemic inflammatory response in COVID-19 patients across the spectrum of respiratory failure severity may help improve patient selection and timing of therapeutics.

In this prospective, observational study spanning the first two years of the SARS-CoV-2 pandemic, we collected longitudinal data in two independent cohorts of inpatients with COVID-19 requiring different levels of respiratory support. We investigated the clinical, biomarker and subphenotype trajectories in COVID-19, examined the prognostic value of host-response profiles on clinical outcomes, and compared trajectories against non-COVID ARDS.

## Methods

Detailed methods are provided in the Supplement.

### Clinical Cohorts

We prospectively enrolled hospitalized patients with COVID-19 in two independent, prospective cohort studies within the UPMC Health System (details in Supplement):

a. **The Acute Lung Injury Registry (ALIR) and Biospecimen Repository** enrolled critically ill COVID-19 patients hospitalized in intensive care units (ICUs)[6].
b. **The COVID INpatient Cohort (COVID-INC)** enrolled moderately ill inpatients with COVID-19 hospitalized in dedicated inpatient wards[11].

### Biospecimen collection

We collected blood samples upon enrollment (baseline – Day 1) and at follow-up time intervals (Days 5 and 10) for those who remained hospitalized, and measured host-response biomarkers.

### Clinical Data Collection

We extracted data on demographics, comorbid conditions, vital signs, and laboratory test results at baseline from the electronic medical record (EMR). We broadly classified baseline respiratory support in four clinical categories, referred to as clinical groups: i) Low Flow Oxygen (LFO), i.e. subjects on conventional nasal cannula or oxygen mask, ii) Non-Invasive Ventilation or High Flow Oxygen (NIV_HFO), i.e. patients either on NIV (continuous or bi-level positive airway pressure) or humidified, heated HFO delivered via nasal cannula or mask, iii) Invasive Mechanical Ventilation (IMV) via endotracheal intubation, and iv) Extracorporeal membrane oxygenation (ECMO) support. We recorded immunomodulatory therapies administered for COVID-19 (steroids, tocilizumab, sarilumab and baricitinib) and timing of administration. We also recorded non-intubated subjects with set limitations in advance care planning with regards to intubation (Do Not Intubate, termed as No-escalation of care). Our primary outcome was 60-day survival from hospital admission, and as secondary outcome we examined respiratory support trajectories starting from date of symptom onset to date of positive PCR testing, hospital/ICU admission, intubation/extubation for mechanically ventilated subjects, discharge, and death.

### Biomarker Measurements

We measured 10 prognostic plasma host-response biomarkers in ARDS/sepsis as previously described (Supplement) but focused our primary analysis on four biomarkers with established relevance in COVID-19 biology: 1. Interleukin-6 (IL-6) [12], a target of approved immunomodulatory therapies for COVID-19, 2. Procalcitonin[13], as a plausible biomarker for secondary bacterial infections, 3. soluble receptor of advanced glycation end products (sRAGE)[14], a biomarker for alveolar epithelial injury, and 4. Angiopoietin-2 (Ang-2) [15, 16], a biomarker for endothelial injury. We included all remaining biomarkers in secondary analyses. From a subset of available plasma samples, we also quantified SARS-CoV-2 RNA with qPCR (i.e. RNA-emia), as previously described.[17, 18]

### Subphenotype Classifications and Statistical Analyses

From available clinical data, we classified patients into inflammatory subphenotypes by applying two biomarker-based parsimonious logistic regression models that had been previously developed via latent class analyses: i) The 4-variable model by Drohan et al. (*“Drohan model”*) utilizing bicarbonate, procalcitonin, sTNFR-1 and Ang-2 levels[7], and ii) the 3-variable model by Sinha et al. (*“Sinha” model*), utilizing bicarbonate, IL-6 and sTNFR-1[19].

We compared continuous and categorical variables between respiratory support groups or subphenotypes with Wilcoxon and Fisher’s test, respectively. We performed log_10_-transformations of biomarker values for statistical analyses. We examined the dynamics of biomarker levels over time using mixed linear regression models against time from hospital admission with random patient intercepts, as well as by comparing biomarker levels between sampling follow-up intervals (Days 1, 5 and 10). For 60-day survival, we constructed Kaplan-Meier curves for time-to-event from hospital admission, as well as Cox proportional hazards models adjusted for age and time from hospital admission. We conducted all analyses in R v4.2.0.

## Results

### Clinical Characteristics of Study Population

Between March 1, 2020 and March 29, 2022, we enrolled a total of 323 patients with COVID-19 **(Table 1)**. Enrolled subjects were predominantly male (57%), white (78%), with median age of 61.4 years. At baseline, we classified subjects into the clinical groups of LFO (n=120, 37%), NIV_HFO (n=92, 27%), IMV (n=88, 27%) and ECMO (n=23, 7%). Patients managed with ECMO were younger, more often white, and had higher BMI than the other clinical groups (Table 1).

**Table 1.**
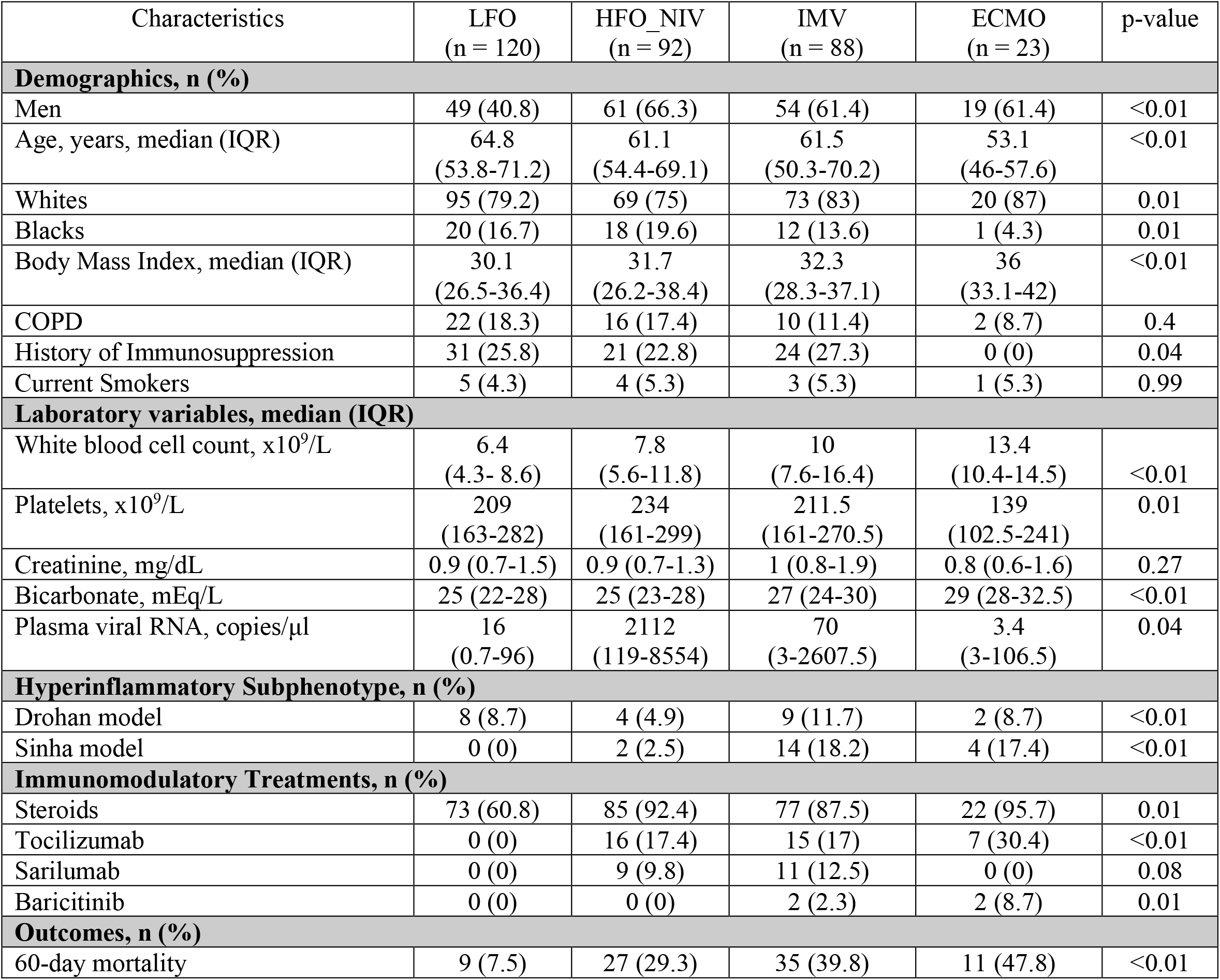
Demographics and Outcomes

| Characteristics | LFO<br>(n = 120) | HFO_NIV<br>(n = 92) | IMV<br>(n = 88) | ECMO<br>(n = 23) | p-value |
| --- | --- | --- | --- | --- | --- |
| **Demographics, n (%)** |  |  |  |  |  |
| Men | 49 (40.8) | 61 (66.3) | 54 (61.4) | 19 (61.4) | <0.01 |
| Age, years, median (IQR) | 64.8<br>(53.8-71.2) | 61.1<br>(54.4-69.1) | 61.5<br>(50.3-70.2) | 53.1<br>(46-57.6) | <0.01 |
| Whites | 95 (79.2) | 69 (75) | 73 (83) | 20 (87) | 0.01 |
| Blacks | 20 (16.7) | 18 (19.6) | 12 (13.6) | 1 (4.3) | 0.01 |
| Body Mass Index, median (IQR) | 30.1<br>(26.5-36.4) | 31.7<br>(26.2-38.4) | 32.3<br>(28.3-37.1) | 36<br>(33.1-42) | <0.01 |
| COPD | 22 (18.3) | 16 (17.4) | 10 (11.4) | 2 (8.7) | 0.4 |
| History of Immunosuppression | 31 (25.8) | 21 (22.8) | 24 (27.3) | 0 (0) | 0.04 |
| Current Smokers | 5 (4.3) | 4 (5.3) | 3 (5.3) | 1 (5.3) | 0.99 |
| **Laboratory variables, median (IQR)** |  |  |  |  |  |
| White blood cell count, x10<sup>9</sup>/L | 6.4<br>(4.3- 8.6) | 7.8<br>(5.6-11.8) | 10<br>(7.6-16.4) | 13.4<br>(10.4-14.5) | <0.01 |
| Platelets, x10<sup>9</sup>/L | 209<br>(163-282) | 234<br>(161-299) | 211.5<br>(161-270.5) | 139<br>(102.5-241) | 0.01 |
| Creatinine, mg/dL | 0.9 (0.7-1.5) | 0.9 (0.7-1.3) | 1 (0.8-1.9) | 0.8 (0.6-1.6) | 0.27 |
| Bicarbonate, mEq/L | 25 (22-28) | 25 (23-28) | 27 (24-30) | 29 (28-32.5) | <0.01 |
| Plasma viral RNA, copies/μl | 16<br>(0.7-96) | 2112<br>(119-8554) | 70<br>(3-2607.5) | 3.4<br>(3-106.5) | 0.04 |
| **Hyperinflammatory Subphenotype, n (%)** |  |  |  |  |  |
| Drohan model | 8 (8.7) | 4 (4.9) | 9 (11.7) | 2 (8.7) | <0.01 |
| Sinha model | 0 (0) | 2 (2.5) | 14 (18.2) | 4 (17.4) | <0.01 |
| **Immunomodulatory Treatments, n (%)** |  |  |  |  |  |
| Steroids | 73 (60.8) | 85 (92.4) | 77 (87.5) | 22 (95.7) | 0.01 |
| Tocilizumab | 0 (0) | 16 (17.4) | 15 (17) | 7 (30.4) | <0.01 |
| Sarilumab | 0 (0) | 9 (9.8) | 11 (12.5) | 0 (0) | 0.08 |
| Baricitinib | 0 (0) | 0 (0) | 2 (2.3) | 2 (8.7) | 0.01 |
| **Outcomes, n (%)** |  |  |  |  |  |
| 60-day mortality | 9 (7.5) | 27 (29.3) | 35 (39.8) | 11 (47.8) | <0.01 |

### SARS-CoV-2 infection timeline and clinical group trajectories

Patients on ECMO had significantly longer time from index COVID-19 qPCR positivity and onset of symptoms, followed by patients on IMV and NIV_HFO, overall indicating later stages of COVID-19 compared to LFO subjects (Figure 1A, Table S1). We examined plasma viral RNA load at time of enrollment and found that NIV_HFO subjects had the highest viral RNA levels (Figure 1B), potentially indicating an earlier phase of SARS-CoV-2 infection with higher viral replication.

**Figure 1:**
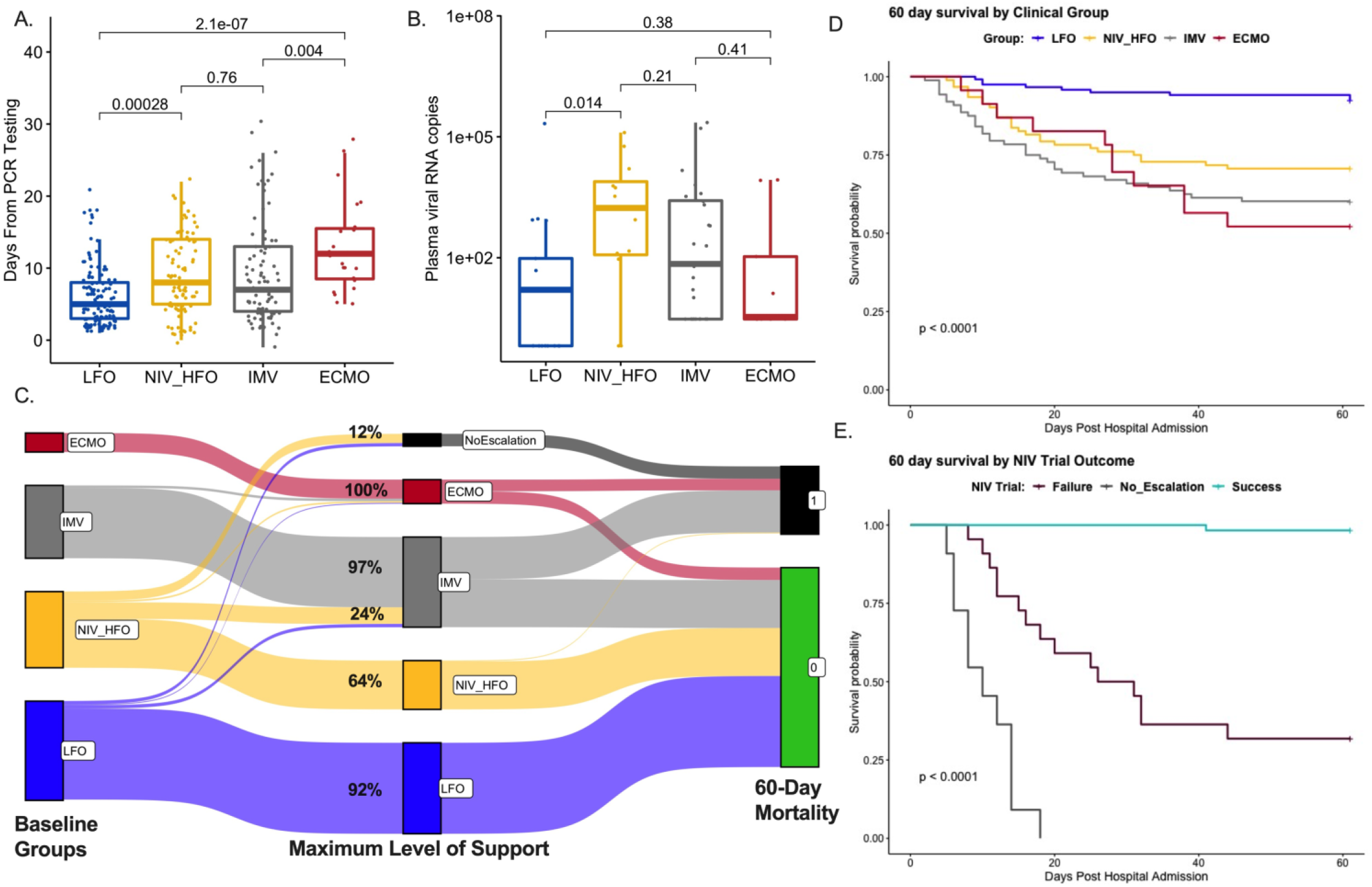
SARS-CoV-2 infection timelines and clinical group trajectories. A. Patients on ECMO had the longest time from the COVID-19 diagnostic qPCR test than other clinical groups. B. Patients on NIV_HFO had the highest levels of plasma viral RNA load (RNA-emia) than the other groups. C. Transition of clinical groups from baseline assignments to the maximum level of respiratory support required during their inpatient stay and then to 60-day outcome (0: survivors, 1: non-survivors). The greatest proportion of transitions occurred in patients on NIV_HFO, with 12% of patients with No-Escalation of care directives transitioning to comfort measures, and 24% requiring escalation to IMV or ECMO. D. 60-day survival curves by Kaplan-Meier analysis for the four clinical groups at baseline. Patients on LFO had markedly improved survival compared to the other three groups. E. 60-day survival curves for NIV_HFO patients at baseline based on the outcome of NIV-HFO trial. All patients with No-escalation of care directives died within 20 days, whereas patients who required escalation to IMV or ECMO had markedly worse survival (67% cumulative 60-day mortality).

Across all groups, we found that at time of enrollment, patients requiring greater respiratory support had worse 60-day survival (Figure 1D). We then examined the clinical group trajectories starting from baseline assignments to maximum level of respiratory support required during hospitalization (Figure 1C). The NIV_HFO group had the most frequent clinical group changes: 12% of patients with No-escalation directives were transitioned to comfort measures and subsequently died, whereas 24% failed a trial of non-invasive support, requiring escalation to IMV or ECMO. This escalation group had markedly worse survival compared to those successfully supported by NIV_HFO (Figure 1E). Patients who failed NIV_HFO were intubated at a median of 7 (2-12) days after admission, whereas patients on IMV at enrollment had been intubated at a median of 3 (0-8) days after admission (p<0.0001), with significantly worse 60-day mortality (67.4%) in these patients with delayed intubation compared to patients on IMV at enrollment (35.2%, p<0.001).

### Plasma Biomarker Trajectories by Clinical Group

In baseline comparisons, IL-6, procalcitonin, and Ang-2 increased with each higher level of support from LFO to ECMO, whereas sRAGE demonstrated the inverse pattern (Figure 2A-D). These observations persisted in both Day 5 and Day 10 comparisons (Figure S1). A similar pattern was seen for many additional biomarkers (Table S2, Figure S2). We constructed mixed linear regression models of biomarker levels from time of admission to address the issue of variability in sampling times by enrollment. This analysis demonstrated declining trajectories for sRAGE levels as a function of time of sampling from hospital admission for patients on NIV_HFO, IMV and ECMO (Figure 2E-H), confirming that sRAGE is a biomarker that peaks earlier in COVID-19 course.

**Figure 2:**
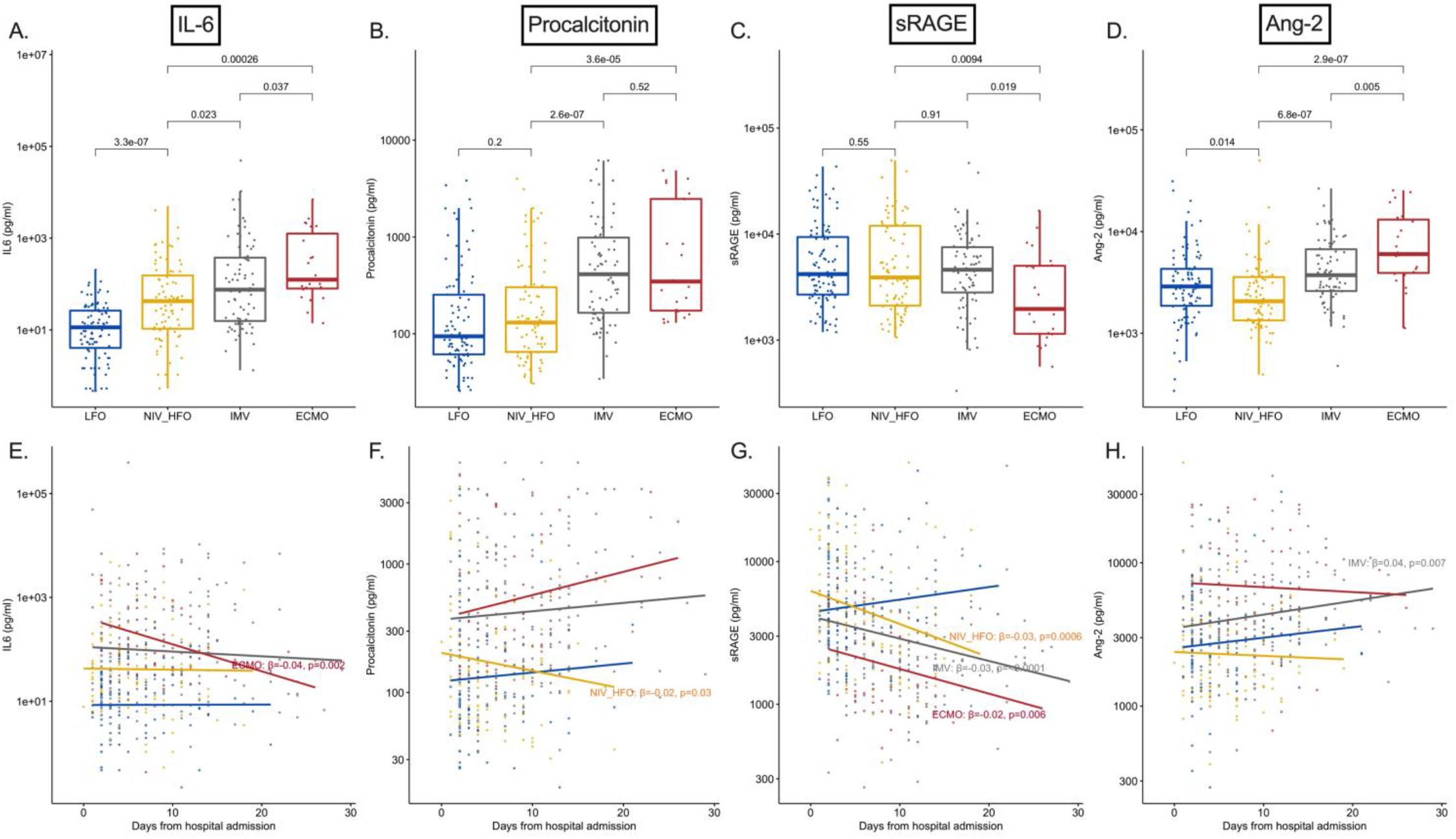
Biomarker trajectories by clinical groups at baseline. A-D: Higher levels of IL-6, procalcitonin and Ang-2 by increasing levels of respiratory support, whereas sRAGE levels were lower in patients on ECMO compared to the other groups. E-H: Trajectories of individual biomarkers in each clinical group as a function of biospecimen sampling time from hospital admission. Statistically significant results (beta co-efficients and p-values) from mixed linear regression models against time from hospital admission with random patient intercepts are displayed. There was a declining trajectory for sRAGE in patients on NIV_HFO, IMV and ECMO.

### Subphenotype Trajectories by Clinical Group

The two parsimonious models (Drohan and Sinha) showed fair agreement in baseline subphenotypic classifications (area under the curve 0.64), with 8% and 7% of subjects classified as hyper-inflammatory, respectively (Figure 3A). Hyper-inflammatory subjects had lower platelets, higher white blood cell count, and worse renal function indices (p<0.01) (Table S3). For subjects with available follow-up biospecimens, subphenotypic classifications from Day 1 to Day 5 were overall stable for the hypoinflammatory subphenotype (with 2% and 8% transitions by the Drohan and Sinha model) but unstable for the Day 1 hyperinflammatory subphenotype with 50% of subjects assigned as hypoinflammatory on Day 5 by both models (Figure 3 and Figure S3).

**Figure 3:**
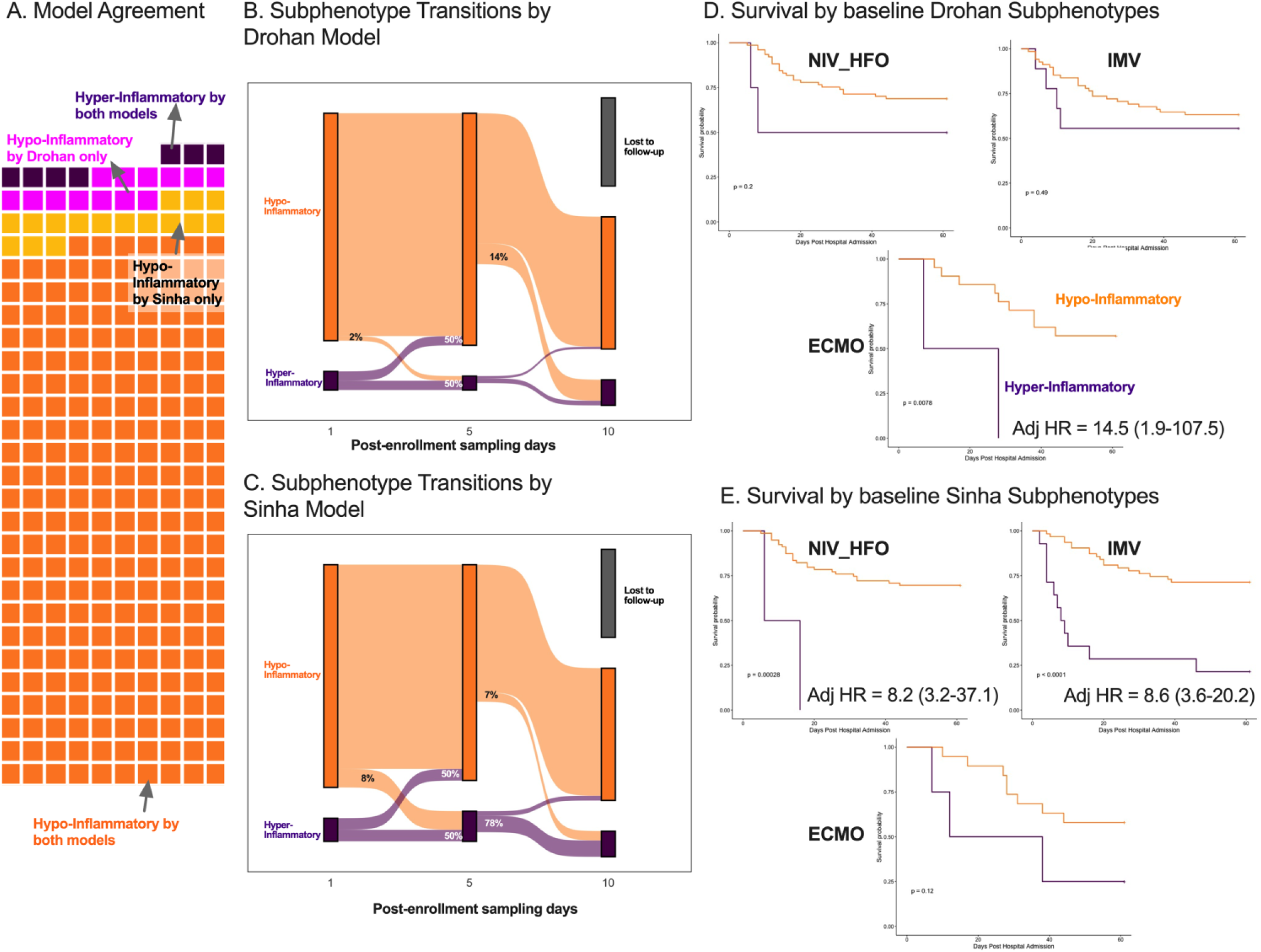
Subphenotypic classifications by parsimonious predictive models, transitions over time, and prediction of outcome. A. Waffle plot for agreement of classifications between the Drohan model (4-variable) vs. the Sinha model (3-variable). Agreement was fair (area under the curve of 0.64). Most subjects (87%) were classified as hypo-inflammatory by both models. B-C. Sankey plot for transition of Drohan and Sinha subphenotypes at each follow-up interval for subjects with available follow-up samples on Day 5. Overall, hypoinflammatory subjects remained stable (2% and 8% transitions, respectively), whereas 50% of hyperinflammatory subjects on Day 1 were classified as hypoinflammatory by Day 5. D-E. Hyper-inflammatory ECMO patients by Drohan model, and hyper-inflammatory NIV_HFO and IMV patients by Sinha model had worse survival in Kaplan-Meier curves and Cox proportional hazards models adjusted for age and time from hospital admission.

### Baseline biomarker levels and subphenotypes prognosticate clinical outcome

We compared baseline biomarker levels between 60-day survivors and non-survivors, stratified by baseline clinical group assignments (Figure 4A-D). The most discriminatory biomarkers for mortality were different amongst clinical groups: sRAGE for LFO and NIV_HFO (p<0.001), and IL-6 for both IMV and ECMO (p<0.01). We also compared biomarker levels between NIV_HFO subjects with successful vs. failed non-invasive support trial, and found that the latter group had significantly higher sRAGE and procalcitonin levels (p<0.0001, Figure 4E-H).

**Figure 4.**
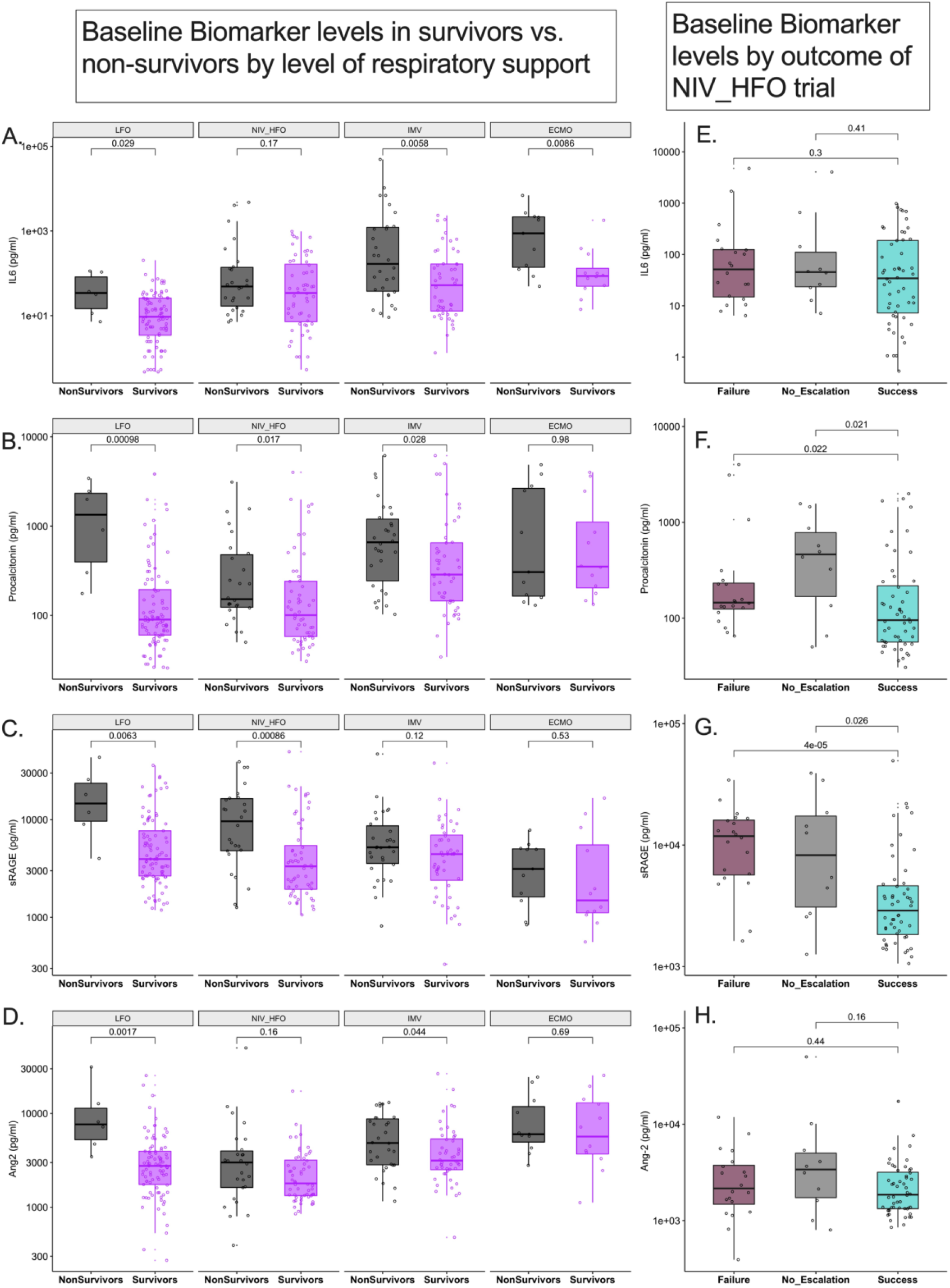
Prognostic effects on 60-day mortality and outcome of NIV_HFO trial by baseline biomarkers. A-D. Higher levels of sRAGE and procalcitonin were associated with LFO and NIV_HFO non-survivors, whereas IL*-*6, procalcitonin, and Ang-2 were associated with IMV non-survivors. IL-6 was the sole discriminatory biomarker in ECMO non-survivors. E-F: Patients with successful trials of NIV_HFO had lower levels of procalcitonin and sRAGE compared to those that failed NIV_HFO trials and went on to require IMV or ECMO.

Baseline subphenotypes by the two models were also predictive of outcome for subjects at different levels of support (Figure 3D-E). By the Drohan model, adjusted and unadjusted survival analysis showed that no hyper-inflammatory ECMO patients survived, whereas by the Sinha model, no hyper-inflammatory NIV_HFO patients survived.

### Immunomodulatory therapies and host-response trajectories

In exploratory analyses, we examined associations between immunomodulatory therapies and biomarker/subphenotype trajectories. As all prescribed therapies were guided by evolving practice guidelines, we sought to mitigate confounding by classifying subjects into three temporally non-overlapping groups who received guideline-congruent treatment at each period (details in the Supplement): i) *no immunomodulation* (first pandemic wave prior to first landmark publication for efficacy of steroids[20]), ii) *steroids only* (from steroids publication to first landmark publication for efficacy of tocilizumab[21]), and iii) *two immunomodulators* (steroids plus tocilizumab, sarilumab or baricitinib). In comparisons of biomarker trajectories following administration of immunomodulatory therapies, we found that receipt of two immunomodulators resulted in markedly higher IL-6 levels compared to those who received only steroids at all sampling intervals (p<0.05, Figure S4). Patients on only steroids or two immunomodulators had significantly lower Ang-2 levels at Day 1 and Day 5 compared to patients on no immunomodulation (Figure S4). Patients on either of the anti-IL-6 agents (sarilumab and tocilizumab) had higher levels of IL-6 and lower levels of Ang-2 compared to the small number of patients on baricitinib (Figure S5), suggesting that anti-IL-6 agents may have accounted for the differences observed between the three treatment groups (Figure S4). Additionally, patients on two immunomodulators had higher frequency of hyper-inflammatory subphenotype by the Sinha model (a model based in part on IL-6 levels) in the baseline interval (Figure S6), likely reflective of the higher IL-6 levels in this patient group.

We identified two subjects for tocilizumab and five subjects for steroids who had samples pre- and post-treatment. We found that tocilizumab administration was associated with higher post-tocilizumab IL-6 levels in both subjects (by 30-fold and 1.5-fold, respectively, Figure S7), whereas post-steroids IL-6 levels on Day 5 were lower than pre-steroids on Day 1 (p=0.04, Figure S8).

## Discussion

We demonstrate distinct clinical and biomarker trajectories that tracked with patient outcomes in a prospective, observational study of hospitalized COVID-19 patients across the spectrum of illness severity and required levels of respiratory support. Host-response biomarker profiling offered prognostic insights, but only when contextualized with the level of respiratory support at time of sampling. A biomarker of alveolar epithelial injury, sRAGE, was predictive of outcome among patients on NIV_HFO, whereas IL-6 carried prognostic value for patients on IMV or ECMO. sRAGE levels declined during hospitalization, whereas other biomarkers showed flat or rising trajectories. Synthesis of host-response profiles with subphenotypic classifications showed an overall low prevalence of the hyper-inflammatory subphenotype in COVID-19 patients, but patients classified to the hyper-inflammatory subphenotype had markedly worse outcomes depending on the level of respiratory support. We demonstrated that immunomodulatory treatments have profound effects on specific biomarker levels, with markedly increased IL-6 in patients receiving anti-IL6 treatments and lower Ang-2 levels in patients on any immunomodulatory treatment.

Our clinical trajectory analyses emphasize the clinical instability of patients enrolled in the NIV_HFO group. Whereas the majority of NIV_HFO were successfully supported without intubation, patients who failed this trial were intubated with considerable delay compared to patients enrolled on IMV, and had the worst mortality of all groups (67%). These findings raise the possibility of inappropriately prolonged non-invasive trials for patients who ultimately required escalation in support, yet at the same time, an approach of early intubation for all would have exposed the “NIV_HFO success” patients to the hazards of IMV. We reveal that patients with a failed NIV_HFO trial had markedly higher sRAGE levels at the time of baseline sampling compared to those with a successful trial (p<0.0001, Figure 4), suggesting that real-time availability of sRAGE levels in conjunction with bedside assessments might help better determine whether continuation of a non-invasive trial is warranted.

The dynamic trajectories of sRAGE levels offer new insights into its prognostic value. As a marker of alveolar epithelial injury, sRAGE levels would be expected to track with COVID-19 severity, yet our analyses showed a seemingly paradoxical pattern, with the sickest patients on ECMO having markedly lower levels. Low sRAGE levels in patients receiving ECMO may simply reflect that such patients receive ultra-protective, low tidal volume ventilation, perhaps mitigating further injury and release of sRAGE into the bloodstream. This hypothesis is further suggested by the extremely high levels of sRAGE in patients who fail NIV_HFO, in whom tidal volumes are difficult to regulate and may induce injurious tidal stretching. Notably, we also found that sRAGE levels consistently decreased over time, a trajectory that was different from the other biomarkers. Given that patients earlier in their course had higher plasma SARS-CoV-2 levels, it is possible that sRAGE may also reflect more active viral replication and lung injury in earlier stages of COVID-19 pneumonia[22]. sRAGE has established prognostic value in non-COVID ARDS as a correlate of radiographic severity, impairments in gas exchange and mechanics, and poor outcome[23, 24]. Our analyses now allow us to view sRAGE not only as a biomarker of disease-related lung injury, but also suggest sRAGE as a potential dynamic metric of patient self-induced or ventilator-induced lung injury.

Biomarker-based subphenotyping with validated models from non-COVID ARDS and respiratory failure cohorts offered prognostic enrichment across the spectrum of COVID-19 severity. Overall, we found low prevalence of the hyper-inflammatory subphenotype (<10%), but when present, the hyper-inflammatory subphenotype carried negative prognostication of striking, and in some subgroups deterministic, strength (e.g. no hyper-inflammatory patient on ECMO survived). Subphenotypic classifications were stable from baseline to middle interval for the hypoinflammatory subphenotype but hyperinflammatory patients demonstrated dynamic transitions, with 50% of them becoming hypoinflammatory on follow-up by both models used for assignments. Prior observations supported stability of subphenotypes in non-COVID ARDS[25], but our data in COVID-19 subjects with acute respiratory failure highlight the need for better understanding of the time-dependent prognostic value and drivers of subphenotypic transitions.

While therapeutic efficacy of steroids and anti-IL6 treatments is well-established[26, 27], the impact of such treatments on host-response biomarkers has not been well studied. Most of the large pragmatic randomized clinical trials for these agents did not involve protocolized biospecimen acquisition for post-hoc biomarker analyses. We leveraged the natural experiment of rapidly evolving clinical practice guidelines at our institution, with time-stamped milestones by practice-changing publications, to infer the impact of different immunomodulatory treatments on measured biomarkers and subphenotypes. Although our analyses cannot control for indication bias or differences in biology from SARS-CoV-2 variants, our results indicate that administered host-targeted therapies in COVID-19 impact prognostic biomarker levels in distinct ways. We found markedly higher IL-6 levels in patients treated with anti-IL6 therapies, consistent with the expected pharmacodynamic effects of tocilizumab and sarilumab binding both the membrane bound and soluble IL-6 receptor, as shown in the COVACTA trial[28]. Conversely, receipt of any immunomodulatory treatment was associated with markedly lower levels of Ang-2, perhaps indicating a protective effect of immunomodulation on endothelial function and vascular biology. Apart from the mechanistic hypotheses that these observations allow, our results also highlight that biomarker-based prognostication using IL-6 or Ang-2 in particular (either in isolation or synthesized in subphenotypes) will be confounded by the effects of immunomodulatory therapies, if blood sampling follows the administered therapy.

Our study has some noteworthy limitations. Our dataset represents a single hospital network, which thus may limit generalizability of our findings, although we enrolled subjects from seven different units and inpatient wards from three different hospitals. There was variability in timing of enrollment due to logistical constraints in obtaining consent from legally authorized patient representatives. Inevitably, some of the biospecimens were obtained later in the hospital course and in most cases after the administration of immunomodulatory treatments. We made concerted efforts to harmonize individual patient trajectories based on objective milestones of COVID-19 illness, such as timings of qPCR testing, symptom onset and hospitalization. Despite the sample size of >300 subjects in our cohort, some of the clinical subgroups were small, and thus cautious interpretation is needed. For practical reasons we merged NIV with HFO subjects in a single group. Therefore, we could not examine for differential effects of spontaneous positive pressure (NIV) vs. negative pressure (HFO) ventilation on host innate immune and injury biomarkers. Additionally, despite our efforts to mitigate confounding by indication in our analyses with the immunomodulatory treatments, these results are only observational and cannot be used for causal inference.

## Conclusions

Longitudinal assessment of the systemic host response in hospitalized COVID-19 patients revealed substantial and predictive inter-individual variability, which was heavily influenced by baseline levels of respiratory support and concurrent immunomodulatory therapies. Future studies examining the predictive value of biomarkers and subphenotypes in COVID-19 and acute respiratory failure need to control for clinical illness trajectory, respiratory support modalities and antecedent immunomodulatory therapies. Robust predictive enrichment with biological subphenotyping of patients considered for enrollment in future clinical trials may allow for better targeting of host modulatory interventions and improved outcomes in critical illness.

### IRB approval and informed consent

We enrolled subjects following admission the hospital and obtained informed consent from the patients or their legally authorized representatives under study protocols STUDY19050099 and STUDY20040036 approved by the University of Pittsburgh Institutional Review Board (IRB).

## Data Availability

All data produced in the present study are available upon reasonable request to the authors

## Acknowledgements

The authors thank the patients and patient families that have enrolled in the cohort studies described in this report. They also thank the physicians, nurses, respiratory therapists, and other staff at the University of Pittsburgh Medical Center (UPMC) Presbyterian/Shadyside Hospitals as well at UPMC East Hospital for assistance with coordination of patient enrollment and collection of patient samples.

## Supplemental Files

### Supplemental File 1: Extended Methods

#### a. The Acute Lung Injury Registry (ALIR) and Biospecimen Repository

ALIR is a prospective cohort study of critically ill patients hospitalized in ICUs at UPMC Presbyterian/Shadyside and UPMC East hospitals. In this study, we included adult patients (18-90 years of age) diagnosed with COVID-19 based on respiratory symptoms, hypoxemia, and confirmatory testing via nasopharyngeal or lower respiratory tract (LRT) quantitative polymerase chain reaction (qPCR) for SARS-CoV-2 RNA. We enrolled subjects following admission to the ICU and obtained informed consent from the patients or their legally authorized representatives under the study protocol STUDY19050099 approved by the University of Pittsburgh Institutional Review Board (IRB). Upon enrollment, we collected blood biospecimens for centrifugation and separation of plasma and other blood constituents, which were stored in −80C until experiments.

#### b. The COVID INpatient Cohort (COVID-INC)

COVID-INC is a prospective cohort study of moderately ill inpatients with COVID-19 hospitalized in dedicated inpatient wards at UPMC Presbyterian/Shadyside hospitals, as well as critically ill patients admitted to the ICU at UPMC East hospital. We enrolled adult patients (18-90 years of age) diagnosed with COVID-19 based on respiratory symptoms, hypoxemia and confirmatory testing via nasopharyngeal or LRT SARS-CoV-2 qPCR. We obtained consent from the patients or their legally authorized representatives under the study protocol STUDY20040036 approved by the University of Pittsburgh IRB. Upon enrollment, we collected blood biospecimens for centrifugation and separation of plasma and other blood constituents, which were stored in −80C until experiments.

##### Biospecimen collection

Upon enrollment (baseline – Day 1), we collected blood biospecimens for centrifugation and separation of plasma and other blood constituents, which were stored in −80C until experiments. We collected follow-up samples on Days 5 and 10 following enrollment for subjects who remained hospitalized, and processed these samples similar to baseline (Day 1) samples.

##### Biomarker measurements

We measured 10 prognostic plasma host-response biomarkers in ARDS/sepsis (interleukins IL-6, IL-8, IL-10, fractalkine, soluble tumor necrosis factor receptor-1 [sTNFR-1], and suppression of tumorigenicity [ST]–2), soluble receptor of advanced glycation end products [sRAGE]), angiopoietin-2 (Ang-2), procalcitonin and pentraxin-3) with a customized Luminex assay (R&D Systems, Minneapolis, MN)[29] as described in our previous studies[7, 30].

### Classification of immunomodulatory therapies

Due to confounding by indication for prescription of immunomodulatory therapies in observational datasets, we focused on the subset of subjects who received guideline-congruent therapies at each phase for the pandemic. Clinical practice guidelines at our institution rapidly evolved based on emerging evidence from randomized clinical trials, and prescriptions were also impacted by availability of each agent (i.e. the case of tocilizumab shortage since August 2021). We therefore defined the following intervals of guideline-based recommendations based on best available evidence in each period for considering the immunomodulatory therapy groups:

1. No immunomodulation: 03/01/2020-06/17/2020: during this period no empiric immunomodulation was advised in our institution, up until the preprint release of the RECOVERY trial results for the efficacy of dexamethasone [20].
2. Steroids only: 06/18/2020-02/11/2021: recommendations for use of steroids only without additional immunomodulators, up until the release of the REMAP-CAP trial results for efficacy of anti-IL6 therapies (tocilizumab and sarilumab) [5].
3. Two immunomodulatory agents (steroids + 2^nd^ agent): 02/12/2021-end of the study. During this period of time for patients who met clinical criteria (escalating respiratory support requirement and plasma levels of CRP >7.5mg/dl), recommendation was to add tocilizumab to steroids at our institution. However, on 08/24/2021 a nationwide shortage of tocilizumab was reported, and guidelines were changed to consideration of sarilumab or baricitinib (publication of the COV-BARRIER study had occurred in the interim) [31]. We considered all subjects who received these immunomodulators in our cohort as eligible based on the clinical criteria deemed by the treating clinicians. We then subdivided this third period of the pandemic into two smaller periods:
  a. Tocilizumab period (02/12/2021-08/24/2021)
  b. Sarilumab/baricitinib period (8/25/2021-end of study).

**Table S1.** Biomarker sampling timeline by respiratory support.

| Characteristics - days,<br>median (IQR) | LFO<br>(n = 120) | HFO_NIV<br>(n = 92) | IMV<br>(n = 88) | ECMO<br>(n = 23) | p-value |
| --- | --- | --- | --- | --- | --- |
| Time from Symptom<br>Onset | 10 (7-13.2) | 12 (7-16) | 12 (8-17) | 17 (11-19) | <0.01 |
| Time from COVID PCR<br>positivity | 5 (3-8) | 8 (5-14) | 7.5 (4-13) | 12 (8.5-15.5) | <0.01 |
| Time from Hospitalization | 3 (2-4) | 4 (2-7) | 4 (2.8-7.2) | 3 (2-5) | 0.01 |
| Time from ICU admission | 2 (-1.2-2) | 2 (1-5) | 3 (2-4) | 3 (2-5) | <0.01 |

**Table S2.** Biomarker levels by respiratory support at baseline.

| Characteristics<br>- median (IQR) | LFO<br>(n = 120) | HFO_NIV<br>(n = 92) | IMV<br>(n = 88) | ECMO<br>(n = 23) | p-value |
| --- | --- | --- | --- | --- | --- |
| IL6, pg/mL | 11.4<br>(4.1-26.3) | 42.3<br>(10.6-153.9) | 75.5<br>(15.7-376) | 124.6<br>(79.8-1340.4) | <0.01 |
| IL8, pg/mL | 12.8<br>(6.2-27.1) | 18.9<br>(12.4-37.9) | 18.9<br>(12.3-29.1) | 29.8<br>(15-59.7) | <0.01 |
| IL10, pg/mL | 6.2 (2.7-7.4) | 6 (1.7-6.6) | 5.6 (1.7- 11.8) | 1.8 (1.5-8.5) | 0.39 |
| Ang2, pg/mL | 2882.3<br>(1859.9-4297.1) | 2064.1<br>(1338-3570.3) | 3725<br>(2599.6-6703.3) | 6003.4<br>(3915.6-13149.4) | <0.01 |
| Procalcitonin,<br>pg/mL | 94.1<br>(61.2-253.3) | 130.4<br>(65-302.5) | 411.9<br>(164.7-987.5) | 346.5<br>(173.3-2473.2) | <0.01 |
| ST2, pg/mL | 73595.6<br>(37377.9-152350.9) | 97230.3<br>(59816.1-171493.5) | 202312.6<br>(120310.3-291827.1) | 202643.7<br>(87070.2-285679.1) | <0.01 |
| Fractalkine,<br>pg/mL | 1491.3<br>(187.2-3014.8) | 2248.6<br>(320.8-3656.8) | 3185.1<br>(1377.6-4845.6) | 3041.2<br>(700.8-4697.7) | <0.01 |
| Pentraxin3,<br>pg/mL | 4403.1<br>(2102.7-11061.2) | 6808.1<br>(4223.3-16992.3) | 12363.7<br>(6538.5-25535.9) | 7826.2<br>(3922.2-14120.8) | <0.01 |
| sRAGE, pg/mL | 4187.1<br>(2684.5-9399.3) | 3895.4<br>(2113.2-11993.2) | 4607.1<br>(2809.4-7525.8) | 1961.5<br>(1146.2-5019.9) | 0.04 |
| sTNFR1,<br>pg/mL | 3465.2<br>(2283.9-5300.3) | 3259.5<br>(2658.1-5077.9) | 4486.2<br>(2951.3-8533.7) | 4946.1<br>(3490-8067.6) | <0.01 |

**Table S3.** Clinical characteristics by baseline subphenotypes.

| Characteristics | Drohan Model |  | p-value | Sinha Model |  | p-value |
| --- | --- | --- | --- | --- | --- | --- |
|  | Hyper-inflammatory<br>(n=23) | Hypo-inflammatory<br>(n=250) |  | Phenotype2<br>(n=20) | Phenotype1<br>(n=253) |  |
| **Demographics, n (%)** |  |  |  |  |  |  |
| Men | 12 (52.2) | 146 (58.4) | 0.72 | 17 (85.0) | 141 (55.7) | 0.02 |
| Age, years, median (IQR) | 60.1<br>(46.6-64.2) | 61.1<br>(53.0-70.3) | 0.25 | 61<br>(55.4-65.8) | 61<br>(52.0-70.0) | 0.97 |
| Race |  |  | 0.89 |  |  | 0.35 |
| White | 19 (82.6) | 196 (78.4) |  | 17 (85.0) | 198 (78.3) |  |
| Black | 4 (17.4) | 42 (16.8) |  | 2 (10.0) | 44 (17.4) |  |
| Other | 0 (0.0) | 12 (4.8) |  | 1 (5.0) | 11 (4.3) |  |
| Body Mass Index, median (IQR) | 29.6<br>(26.9-34.1) | 31.9<br>(27.0-37.7) | 0.31 | 33.7<br>(29.0-37.1) | 31.6<br>(26.6-37.1) | 0.52 |
| COPD | 3 (13.0) | 41 (16.4) | 0.9 | 1 (5.0) | 43 (17.0) | 0.28 |
| Immunosuppression | 6 (26.1) | 55 (22.0) | 0.85 | 5 (25.0) | 56 (22.1) | 0.99 |
| Current Smokers | 3 (13.0) | 9 (3.6) | 0.11 | 1 (5.0) | 11 (4.3) | 1 |
| **Laboratory variables, median (IQR)** |  |  |  |  |  |  |
| White blood cell count, x10<sup>9</sup>/L | 11.7<br>(6.4-19.9) | 8.1<br>(5.4-11.6) | 0.05 | 13.6<br>(8.9-24.2) | 7.8<br>(5.4-11.4) | <0.01 |
| Hemoglobin, g/dL | 9.2 (8.2-10.6) | 11.9 (10.4-13.5) | <0.01 | 11.0 (9.4-13.4) | 11.8 (10.2-13.4) | 0.33 |
| Platelets, x10<sup>9</sup>/L | 169.0<br>(124.0-201.0) | 213.0<br>(156.0-277.5) | 0.02 | 124.0<br>(96.5-198.0) | 210.0<br>(160.0-278.0) | <0.01 |
| Creatinine, mg/dL | 3.6 (3.2-7.0) | 0.9 (0.7-1.3) | <0.01 | 2.0 (1.1-2.9) | 0.9 (0.7-1.4) | <0.01 |
| Bicarbonate, mEq/L | 24.0<br>(23.0-27.0) | 26.0<br>(23.0-29.0) | 0.09 | 25.0<br>(22.8-27.0) | 26.0<br>(23.0-29.0) | 0.11 |
| Plasma viral RNA, copies/ul | 3.0<br>(3.0-80.6) | 60.6<br>(3.0-2040.0) | 0.29 | 65.3<br>(17.4-6245.5) | 32.0<br>(3.0-918.0) | 0.54 |
| **Immunomodulatory Treatments, n (%)** |  |  |  |  |  |  |
| Steroids | 19 (82.6) | 204 (81.6) | 1 | 18 (90.0) | 205 (81.0) | 0.48 |
| Tocilizumab | 4 (17.4) | 30 (12.0) | 0.67 | 9 (45.0) | 25 (9.9) | <0.01 |
| Sarilumab | 0 (0.0) | 18 (45.0) | 0.36 | 4 (57.1) | 14 (38.9) | 0.63 |
| Baricitinib | 0 (0.0) | 3 (7.5) | 1 | 0 (0.0) | 3 (8.3) | 1 |
| **Outcomes, n (%)** |  |  |  |  |  |  |
| 60-day mortality | 11 (47.8) | 60 (24.0) | 0.02 | 16 (80.0) | 55 (21.7) | <0.01 |

**Figure S1.**
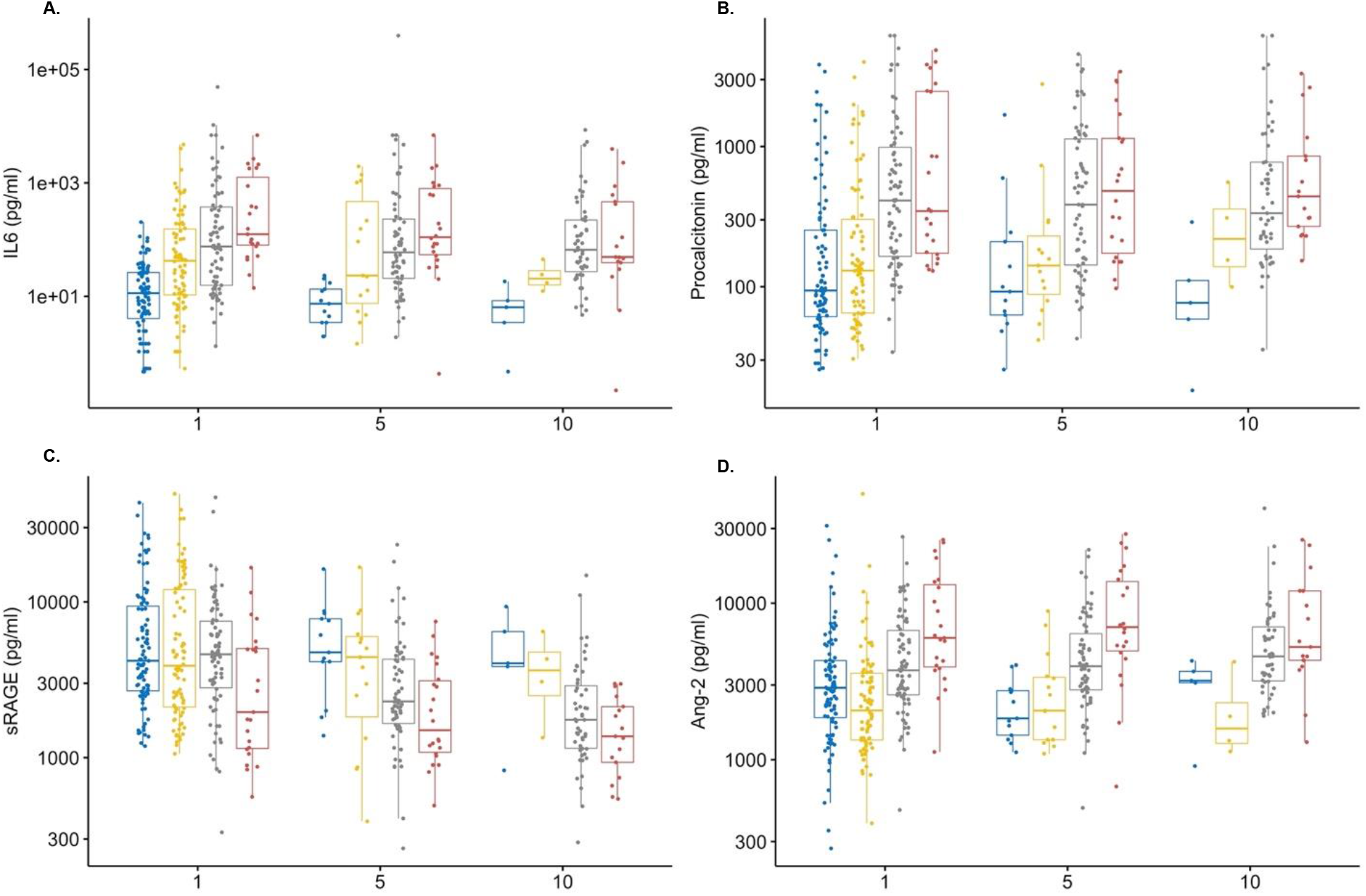
Trajectories of the four primary biomarkers of interest by respiratory support levels. A-D: Higher levels of IL-6, procalcitonin and Ang-2 by increasing levels of respiratory support, whereas sRAGE levels were lower in patients on ECMO compared to the other groups. This pattern proved persistent throughout all sampling intervals.

**Figure S2.**
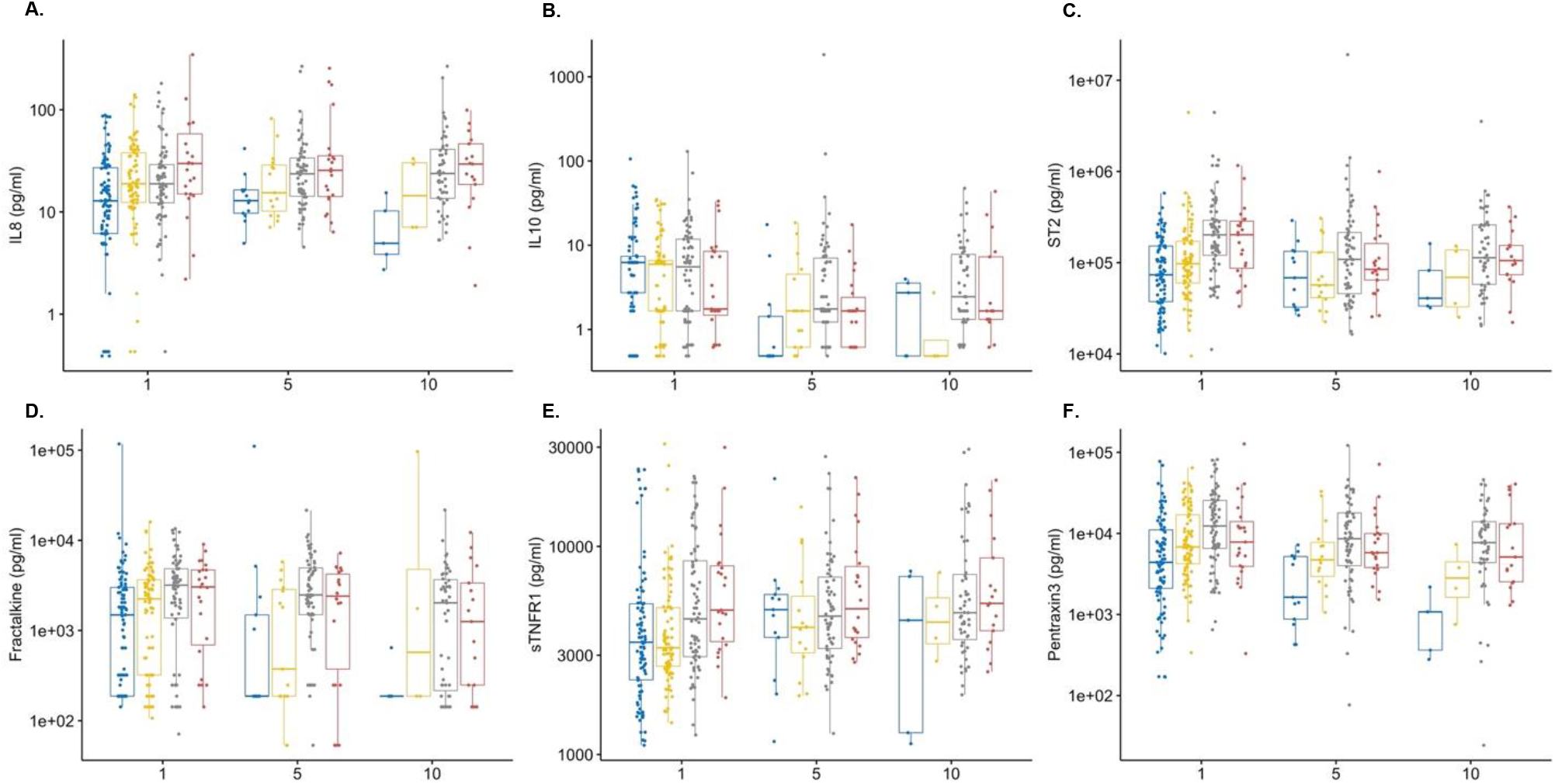
Trajectories of six biomarkers by respiratory support (secondary analyses). A-D: Higher levels of IL-8, ST2, fractalkine, sTNFR1, and pentraxin-3 by increasing levels of respiratory support.

**Figure S3:**
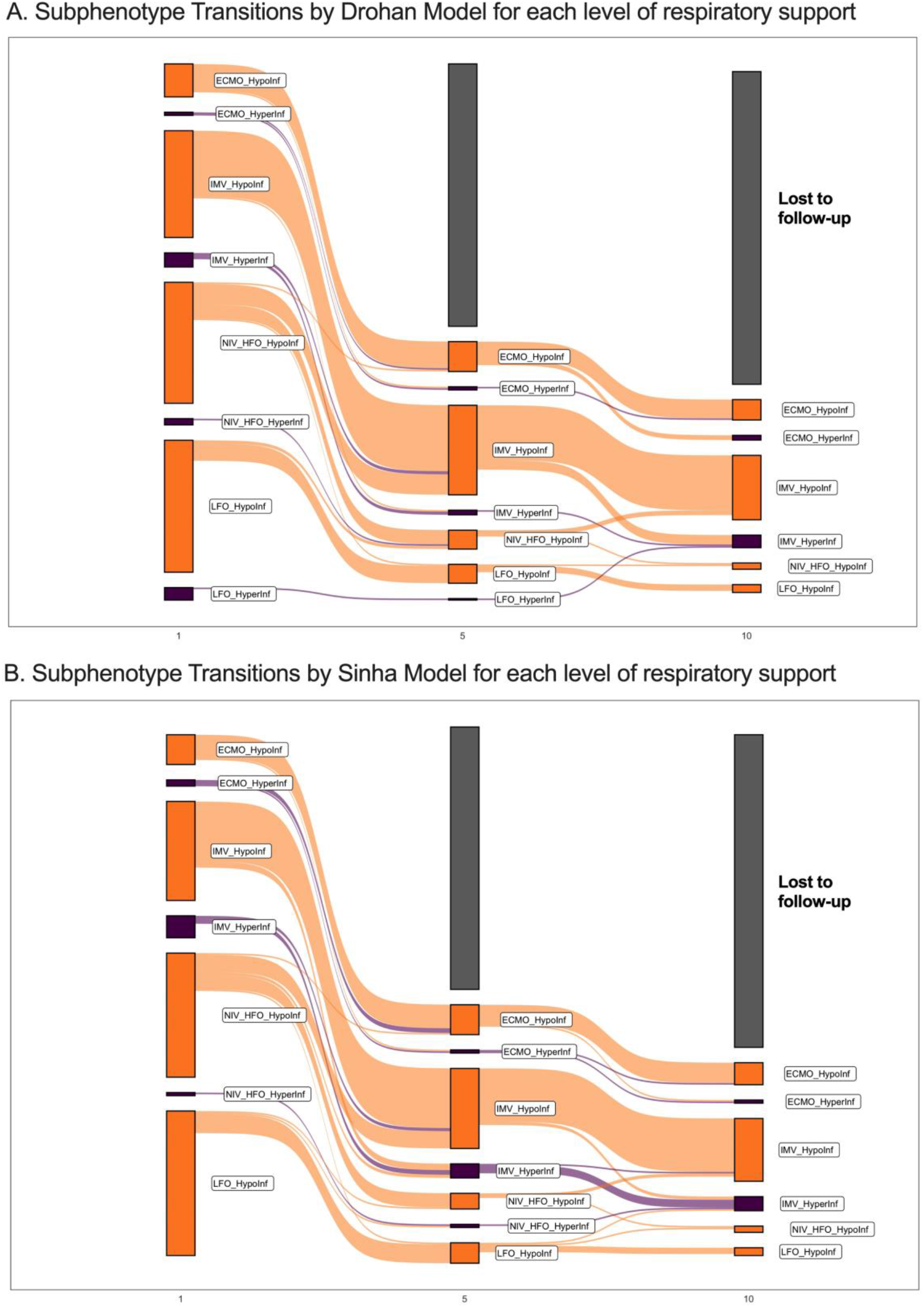
Subphenotypic classifications and transitions over time. Sankey plot for transition of Drohan (A) and Sinha (B) subphenotypes at each follow-up interval by clinical group.

**Figure S4.**
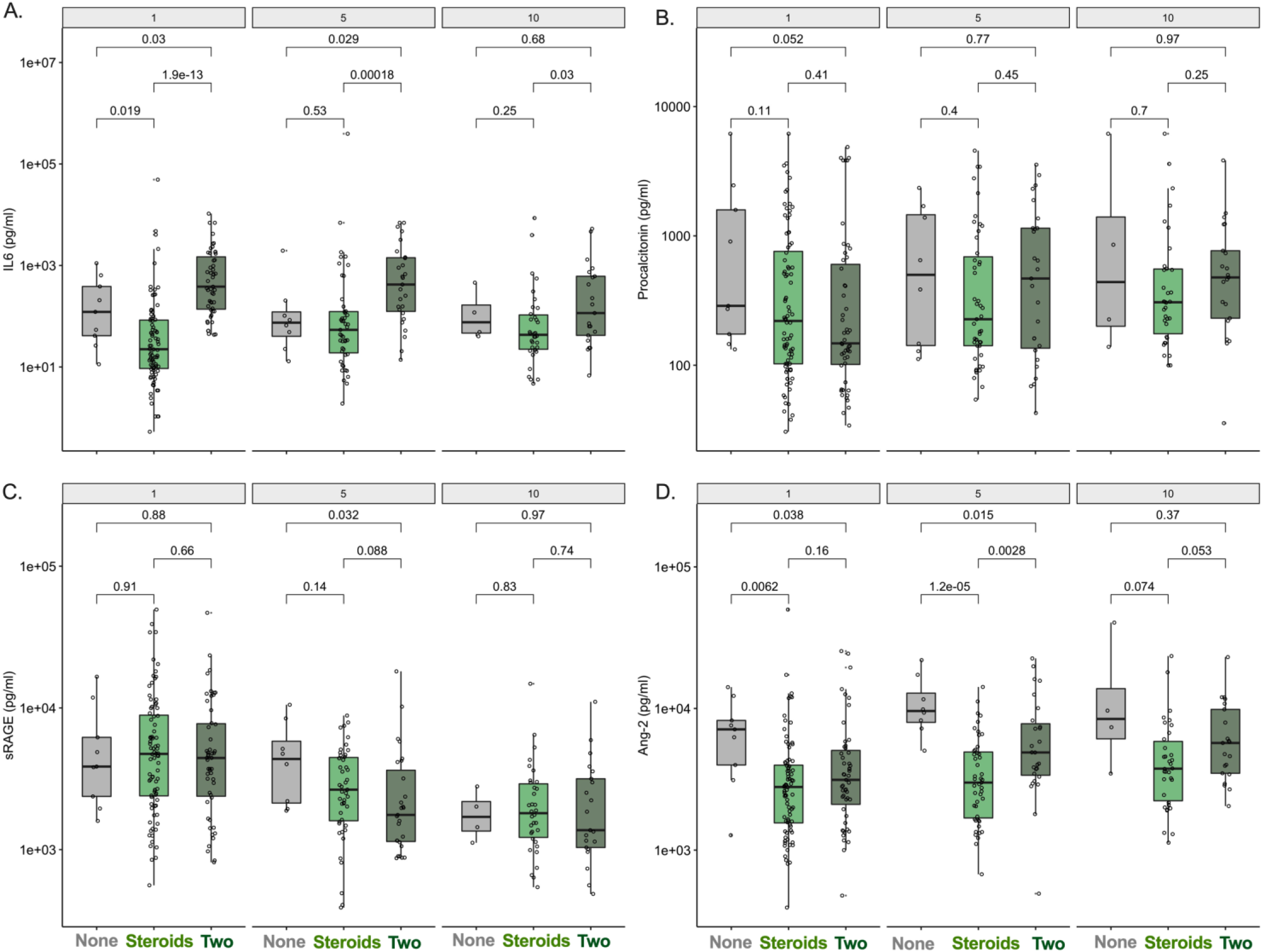
Biomarker trajectories by immunomodulatory therapies. A. Patients who received two immunomodulators (steroids and anti-IL6 therapy) had significantly elevated IL-6 levels at baseline and all follow-up intervals. B-C. Immunomodulation had no significant effects in procalcitonin and sRAGE levels. D. Patients who receive one or more immunomodulatory agents had lower Ang-2 levels on Day 1 and Day 5.

**Figure S5.**
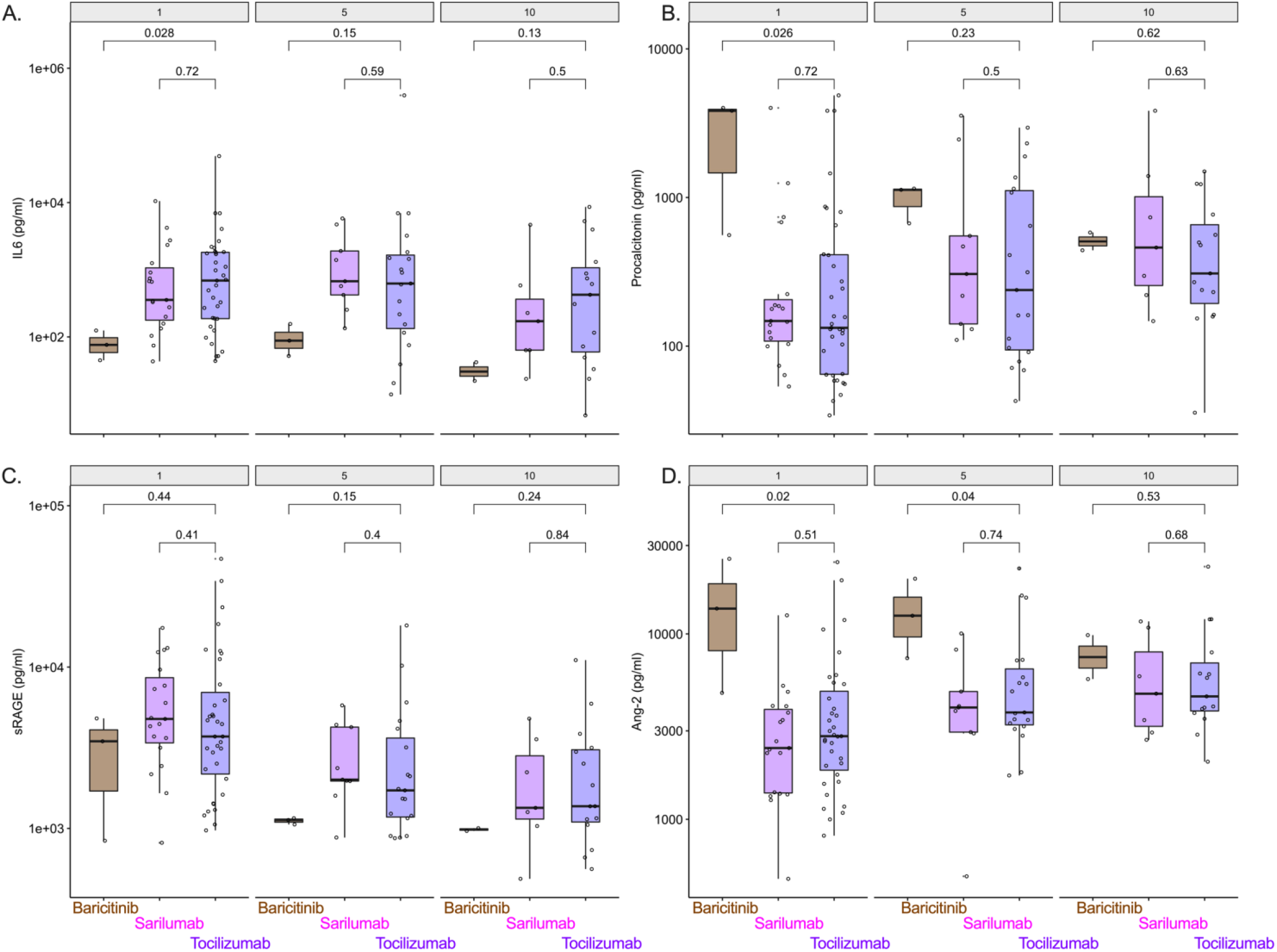
Biomarker trajectories by anti-IL6 therapy (sarilumab or tocilizumab) and baricitinib. A-D: Patients receiving IL-6 receptor antagonists had higher levels of IL-6 and lower levels of Ang-2 and procalcitonin compared to patients receiving barcitinib.

**Figure S6.**
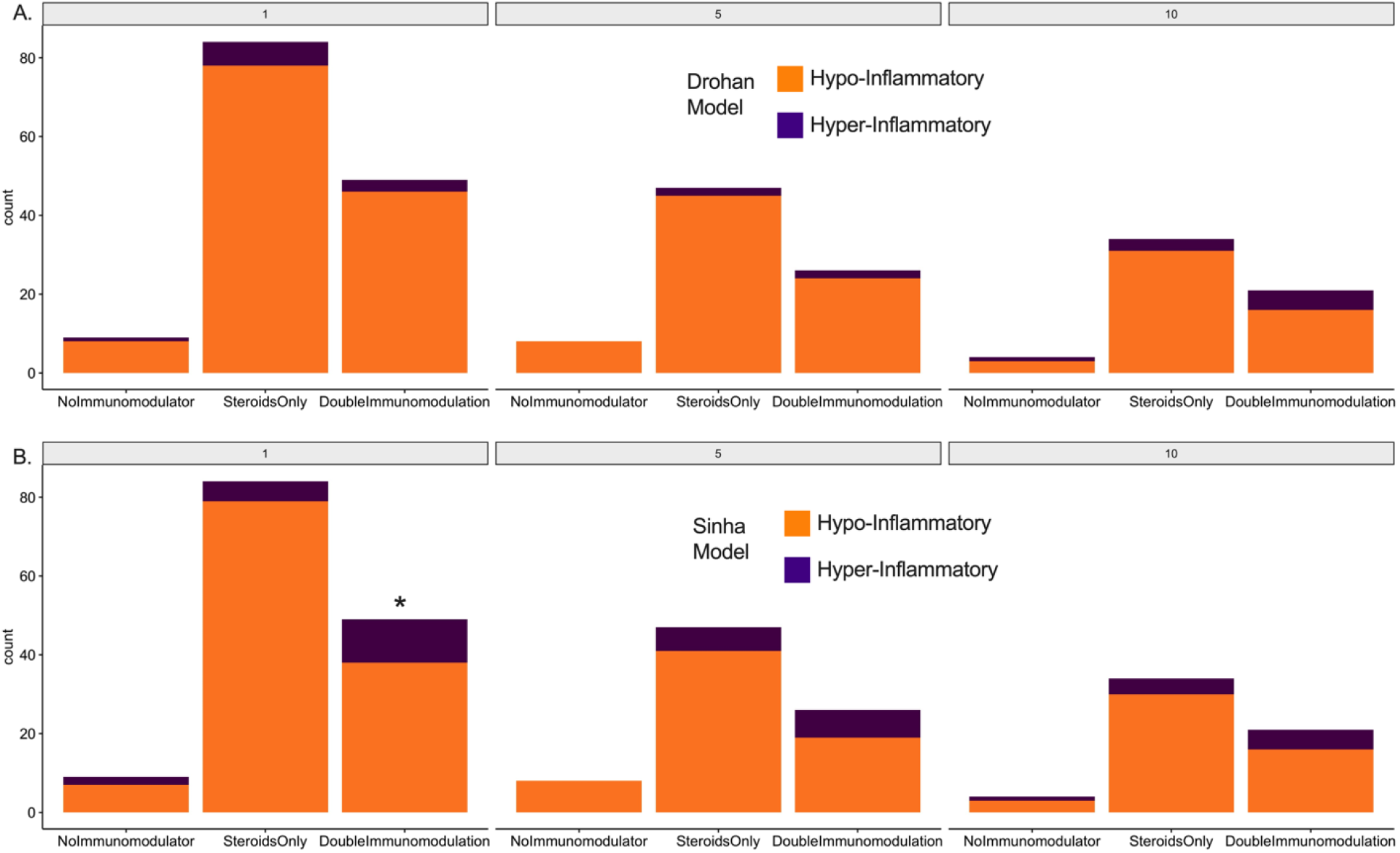
Distribution of subphenotypes by the Drohan model (A) and the Sinha model (B), stratified by immunomodulatory therapies during the three timepoints of follow-up. We found significantly higher proportion of the hyper-inflammatory subphenotype by the Sinha model in patients with two immunomodulatory treatments in the baseline interval (p<0.05).

**Figure S7:**
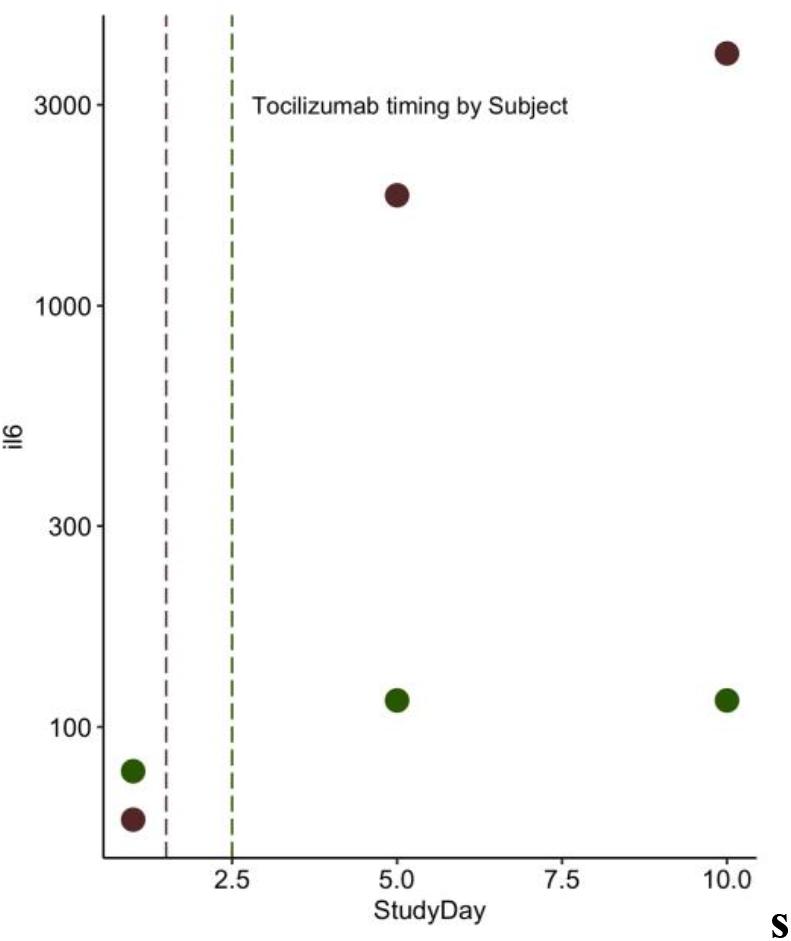
Pre and post-tocilizumab levels for 2 subjects with available biospecimens. Patients (n=2) with serum samples pre- and post-tocilizumab therapy have higher levels of IL-6 after tocilizumab administration.

**Figure S8.**
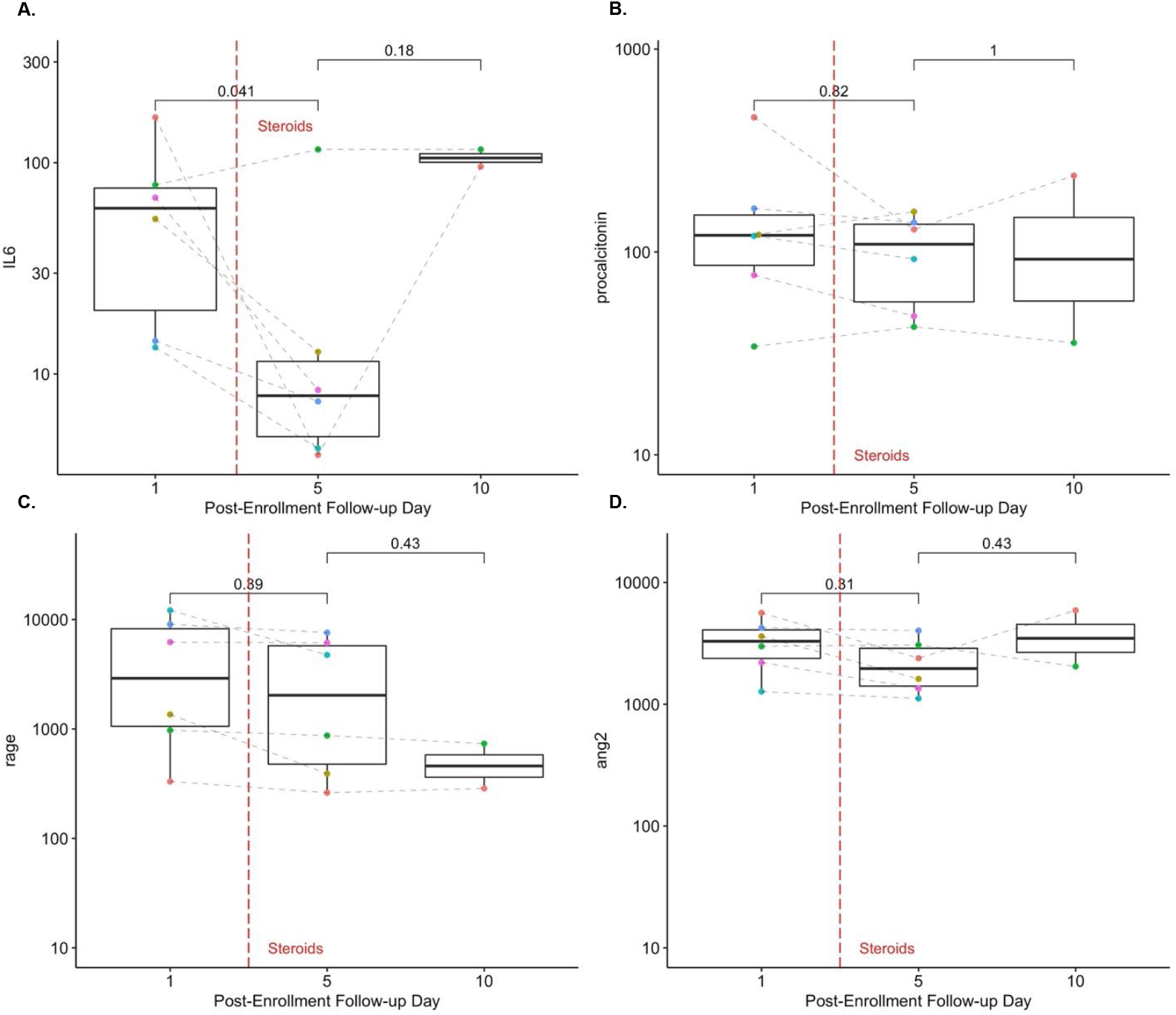
Pre- and post-steroids biomarker levels. Patients (n=5) with serum samples pre- and post-steroids therapy have lower levels of IL-6 on Day 5 following steroid administration.

